# Latent tuberculosis infection diagnosis and management in pediatric primary care: The role of electronic health record-based screening

**DOI:** 10.64898/2026.01.18.26344307

**Authors:** Julia Fink, William B Burrough, Mariamawit Tamerat, Shereen S. Katrak, Tessa Mochizuki, Katya Salcedo, Amit Chitnis, Charlotte Hsieh, Zarin Noor, Gena Lewis, Devan Jaganath

## Abstract

**Background and Objectives:** Annual tuberculosis (TB) screening is recommended for all children and adolescents in the United States, but gaps remain in the diagnosis and treatment of latent TB infection (LTBI). We utilized the electronic health record (EHR) to examine the pediatric LTBI care cascade and assessed if an EHR note template could increase risk factor screening.

**Methods:** We extracted EHR data from well-child and -adolescent visits at a federally qualified health center in Northern California from 2014-2020. We constructed the LTBI care cascade from screening through treatment and performed multivariable logistic regression to assess factors associated with completion of cascade steps. A TB risk factor question was added to the progress note template in 2014, and we measured the change in TB risk factor screening and testing over time.

**Results:** We included 10,409 children from 18,681 visits, with median age of 6.9 years (IQR 2.8-12.1). Most visits (90%) had completed risk factor screening, and the note template significantly increased screening over time. However, 20% with a TB risk factor had testing ordered, though the proportion increased from 7% to 33% throughout the period. Of those tested, 4% had a positive test, and the majority completed subsequent steps. Children under 5 years old were more likely to have risk factor screening than older children but were less likely to be tested.

**Conclusions:** LTBI risk factor screening is high, but ongoing gaps in testing could have led to underdiagnosis. Simple EHR-based solutions have the potential to improve pediatric TB care.

**Article Summary:** We examined the care cascade for latent tuberculosis infection (LTBI) in children and evaluated the role of the electronic health record to support LTBI care.

**What’s Known on This Subject:** The American Academy of Pediatrics recommends annual tuberculosis screening at well-child visits, but gaps remain in completion of the care cascade. Electronic health record (EHR) systems can be utilized to monitor and guide completion of health maintenance activities.

**What This Study Adds:** Prior efforts to characterize the latent tuberculosis infection (LTBI) care cascade for children were incomplete, with limited EHR-based interventions. This study provides a full assessment of the LTBI care cascade and examines the role of the EHR to improve screening.

## INTRODUCTION

In the United States (US), tuberculosis (TB) disease remains a significant public health challenge. After decades of decreasing incidence, TB disease cases have increased in the US since 2021.^1,2^ To reverse this trend, it is critical to identify and treat latent tuberculosis infections (LTBI), as approximately 80% of TB in the US is attributable to LTBI reactivation and progression to disease.^3^

Preventing TB disease in children is critical. After exposure, children have a higher risk of developing a primary disease compared to adults, and if disease develops, they are at higher risk of more severe disease including TB meningitis.^4–6^ Moreover, children with LTBI have a longer lifetime risk for reactivation than adults and thus greater potential benefit of treatment.^7^ Children tolerate LTBI treatment medications better than adults, with lower rates of adverse events and treatment interruption.^8,9^ Consequently, the American Academy of Pediatrics (AAP) recommends annual screening for risk factors associated with TB and subsequent treatment of those with evidence of LTBI.^10^

The steps needed from screening to treatment are referred to as the LTBI care cascade. Among the limited studies focusing on this LTBI care cascade in children, there are significant gaps that result in missed opportunities for treatment.^11^ There is a critical need to better characterize the pediatric LTBI care cascade and examine if tools in the electronic health record (EHR) could aid in closing any gaps. Therefore, we characterized the LTBI care cascade over seven years at a pediatric Federally Qualified Health Center (FQHC) in Northern California and analyzed the impact of integrating TB risk factor screening into the EHR well-child and -adolescent note templates.

## METHODS

### Patient population and Study Design

We performed a retrospective cohort analysis of patient visits at a pediatric, primary care FQHC encompassing four clinical sites located in Oakland, Alameda County, CA, US. These clinics see children and adolescents from birth through 24 years old and serve patients regardless of insurance status, ability to pay and immigration or citizenship status.^12^ Alameda County itself is a highly diverse region, with approximately 33% of its population born outside the US.^13^ Notably, this county has the 4th highest case rate of TB for all jurisdictions in California, and 1.4 times higher than the California state rate.^14^ Our retrospective cohort was created through chart extraction of visits occurring during March 2014 through February 2020.

### Data Collection

All well-child and -adolescent visits were extracted from the FQHC clinic group EHR software Epic (Epic Systems, Verona, WI). For these visits, patient demographic information (age, sex, self-reported race, and self-selected preferred language), visits diagnosis and encounter orders were obtained. Starting in 2014, the well-child note template used by providers included a section for documentation of new TB risk factors since last visit. This section contained options for four TB risk factors as guided by the California Department of Public Health (CDPH) Pediatric TB Risk Assessment: 1) birth in a country with elevated TB rate; 2) travel or residence in a country with elevated TB rate for at least 1 month; 3) immunosuppression, current or planned; and 4) close contact with someone with infectious TB disease.^15^ The LTBI risk factor drop down list and the 100 characters following this section were also extracted to capture any clarifying free text inserted by providers. Laboratory, imaging, and medication orders were obtained from medical records for patients identified with LTBI risk factors.

### Cohort Creation

We included all visits for children aged 1-18 years old with encounter diagnoses listed in **Supplemental Table 1**. Visits were excluded if the patient had a prior diagnosis or treatment for TB disease or LTBI based on encounter codes, free text and ICD 9 or 10 codes, with confirmation by chart review.

### LTBI Care Cascade

The LTBI care cascade consists of a) screening for TB risk factors; b) testing those with risk factors for TB exposure with tuberculin skin testing (TST) or an interferon gamma release assay (IGRA); c) ruling out TB disease with a physical exam and chest X-ray (CXR); d) initiation of LTBI treatment; and e) completion of LTBI treatment.^16^ We define each step in the **Supplemental Methods**. At this clinic, QuantiFERON-TB Gold Plus (QFT, QIAGEN, Germantown, MD) was the available IGRA.

### Statistical Analysis

We described the cohort using summary statistics. The first step of the care cascade was the proportion of the total patient visits where TB risk factor screening was performed. For each subsequent step, the proportion that completed that step was calculated. We constructed a care cascade of the total population and stratified by age group and those with a non-English language preference. Annual changes in risk factor screening rates were assessed using logistic regression, and subgroup comparisons were performed using chi-squared testing. We assessed the demographic factors associated with completion of a given LTBI care cascade step using the Wilcox signed-rank test for continuous variables and a chi-squared or Fisher’s exact for categorical variables. Significance was defined as a p-value < 0.05. Multivariate logistic regression models were constructed utilizing demographic variables, TB risk factor and, when applicable, a TB infection test. As patients had multiple visits over the period of study, we adjusted for clustering by patient, and we reported an adjusted odds ratios (aORs) with a 95% confidence interval (CI) based on robust standard errors. RStudio 2022.07.1 was used to conduct this analysis with *lubridate, dplyr, readxl, stats, aod, clubSandwich and stringr* packages (Posit PBC, Boston, Massachusetts, USA).

### Ethical Review

This study was reviewed and approved by Institutional Review Board (IRB) of the University of California San Francisco.

## RESULTS

### Study Population

From March 2014 through February 2020, we identified 18,681 patient visits meeting inclusion criteria, representing 10,409 children and adolescents **(Figure 1)**. Patient-level demographic characteristics are presented in **Table 1**. Most patients had their first clinic visit before the age of 12. Among those with documented information on race, the highest self-identified race was Black or African American (63%), followed by White (7%), Asian (7%) or multi-racial (4%); for ethnicity, 24% of patients identified as Hispanic. In total, 19% of patients preferred a language other than English. Most patients utilized publicly funded health insurance.

**Figure 1.**
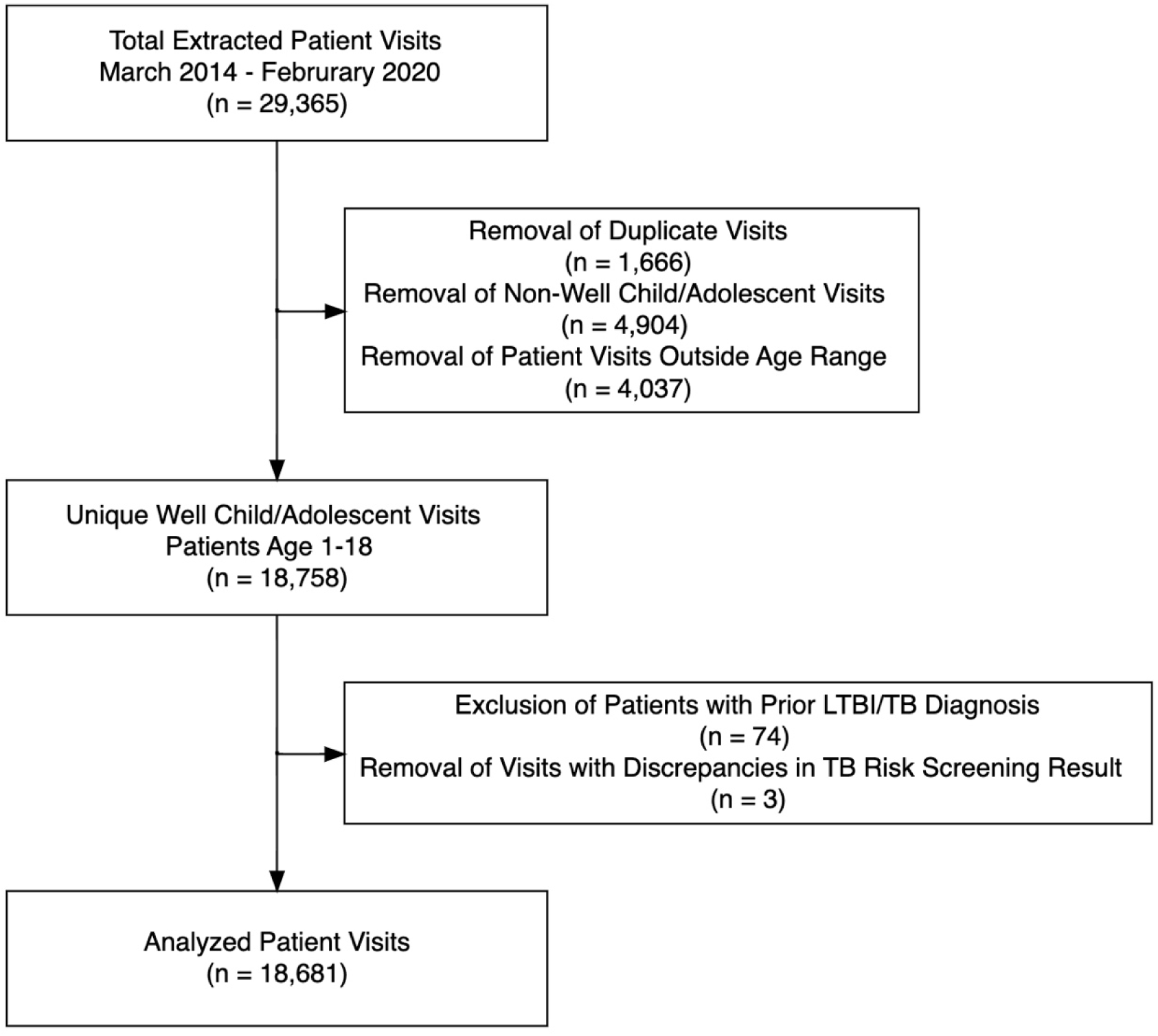
Cohort Flow Diagram. **Alt Text:** Flowchart showing the number of visits that were analyzed after removal due to duplicates or not meeting inclusion criteria. Ultimately, 18,681 well-visits were included from 10,409 patients.

**Table 1.** Descriptive characteristics of cohort. Alt Text. A summary table of the demographic characteristics of the patients included in the study. Overall, the median age was 6.9 years, half female, and 19% preferred a language other than English.

| **Demographic Characteristics** | **Unique Patients<sup>1</sup>**<br>**(n=10,409)** |
| --- | --- |
| **Age at first visit in years** (median, IQR) | 6.9 (2.8, 12.1) |
| **Age category at first visit in years** (N, %) |  |
| Age 1 to 4 | 4,051 (38.9) |
| Age 5 to 11 | 3,683 (35.4) |
| Age 12+ | 2,675 (25.7) |
| **Sex** (N, %) |  |
| Female | 5,121 (49.2) |
| **Race** (N, %) | (n= 10,238) |
| Reported | 6,206 (60.6) |
| Asian | 447 (7.2) |
| Black/African American | 3,921 (63.2) |
| Multi-Racial | 268 (4.3) |
| Native American /Alaskan Native | 10 (0.2) |
| Pacific Islander/Native Hawaiian | 72 (1.2) |
| White/Caucasian | 456 (7.3) |
| Other | 4,032 (39.4) |
| **Ethnicity<sup>2</sup>** (N, %) | (n= 9,313) |
| Hispanic | 2,309 (24.8) |
| **Preferred Language** (N, %) |  |
| English | 8,408 (80.8) |
| Spanish | 1,128 (10.8) |
| Arabic | 326 (3.1) |
| Tigrinya/Tigre | 152 (1.5) |
| Other | 395 (3.8) |
| **Insurance** (N, %) | (n = 10,372) |
| Public | 9,571 (92.3) |
| **Number of Visits per Patient** (median, IQR) | 1 (1, 2) |

Abbreviation: IQR – interquartile range
1. Population total is 10,409 for unique patients unless otherwise indicated.
2. Not-imputed, only analyzed patient if selected “Hispanic or Latino” or “Not Hispanic or
Latino”

### Care Cascade

Providers documented completion of TB Risk Factor assessment for 16,838 (90%) visits (**Figure 2**), and a new risk factor was identified in 1,277 (8%) visits. The most common risk factor identified was related to country of birth (391/1,277, 31%), followed by travel or previous residence in a country with an elevated TB rate (341/1,277, 27%).

**Figure 2.**
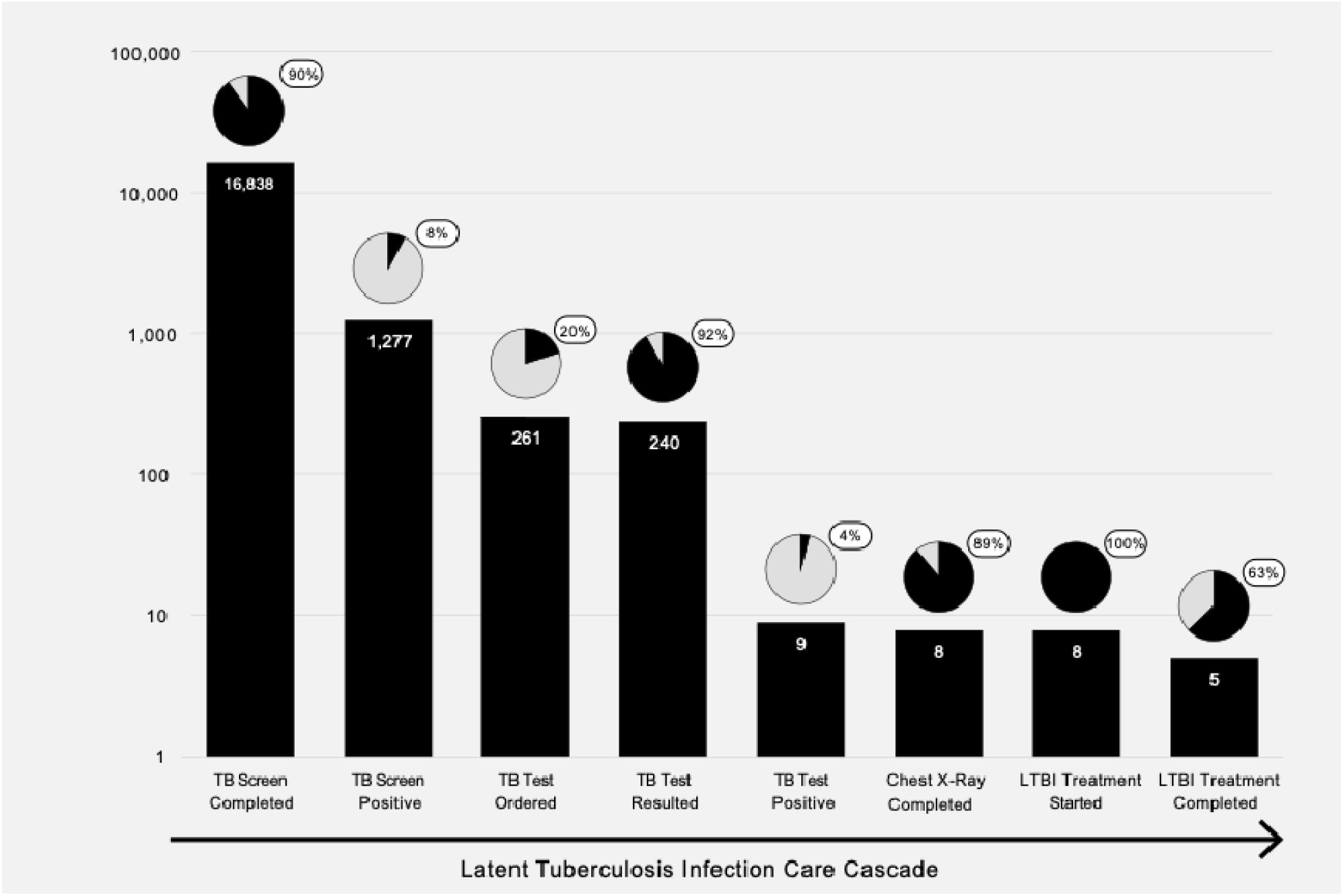
Care cascade for pediatric TB infection (N=18,681 visits). Each bar represents the total number of visits where the step was completed or result was positive, and is the denominator for the next step. The pie chart above each bar indicates the percentage completed or positive. **Alt Text:** Bar chart showing each step of the care cascade on the x-axis, with the number where a given step was completed or result was positive on the y-axis. A pie chart is above each bar that shows the percentage who completed each step or had a positive result.

Only 20% of patient visits (n = 261) with a new TB risk factor had an associated QFT or TST ordered, of which 92% (n = 240) of tests were completed. In total, 9 patients (4%) had positive diagnostic tests, indicating TB infection. One of these patients was TST positive but had previously received a Bacille Calmette-Guérin (BCG) vaccination and repeat QFT testing was negative. All other patients (8) with a positive test result had a CXR ordered, completed and interpreted as not consistent with TB disease. They were all started on LTBI treatment, and five (63%) of these eight patients had subsequent documentation of completion of LTBI treatment. Similar trends were seen when care cascades were developed by age group and among non-English speakers **(Supplemental Figures 1-4**); however, risk factor screening was lower in adolescents 12 years and older (77%) though testing was higher (28%).

### Screening and Testing over Time

Screening was statistically significantly more likely to occur during patient visits in 2015 compared to 2014 (76% in 2014 versus 92% in 2015 p < 0.01, **Figure 3A**). This association remained when adjusted for age group, sex, language and race (aOR 3.98, 95% CI 3.07-4.91). From 2015-2020, the proportion of visits with risk factor screening remained high (greater than 90%), although risk factor screening rates for adolescents 12-19 years remained below 90% throughout the study period (**Supplemental Figure 5)**. For patient visits where a TB risk factor was identified, rate of diagnostic test ordering was initially 7% (95% CI 4-12) in 2014 and 3% (95% CI 1-8) in 2015 (**Figure 3B**), but significantly increased to 10% by 2016 (2015 vs. 2016, aOR 3.36, 95% CI 1.08-10.50) and by 2020 was 33% (95% CI 21-47). These trends were also seen across age group categories (**Supplemental Figure 6**), though again children under five years underwent less testing.

**Figure 3.**
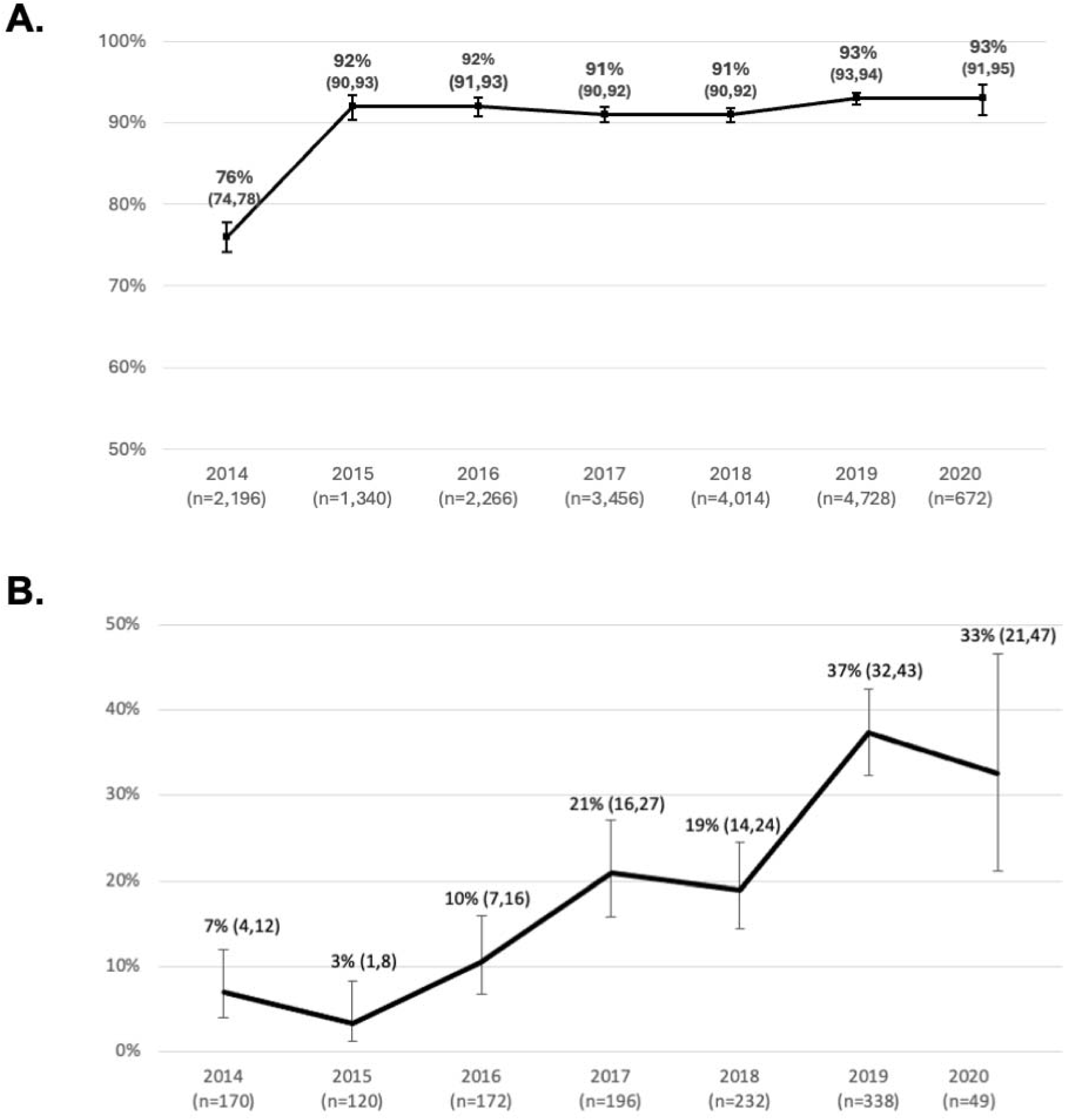
TB infection risk factor screening and testing over time. A) Percentage of well visits with TB risk factor screening by year with 95% confidence interval (CI). B) Percentage of patients with positive TB risk factor screening that had a TB infection test (Tuberculin Skin Test or QuantiFERON Gold) ordered by year with 95% CI. **Alt Text**. Line graphs that show the percentage who were screened and tested for tuberculosis infection by year.

### Multivariable factors associated with screening and testing for pediatric TB infection

Factors associated with TB screening and testing in the univariate analysis can be found in **Supplemental Tables 2-3**. In multivariable analysis (**Table 2**), we found that children younger than five years were significantly more likely to be assessed for risk factors than older age groups but were also less likely to be tested. Additionally, in visits for non-English speaking patients, rates of screening and testing were higher compared to similar English-speaking patients. Those with risk factors that were not reported or not part of the CDPH Pediatric TB Risk Assessment were less likely to be tested compared to those with a risk factor of birth in a country with an elevated TB rate.

**Table 2.** Multivariable analysis of factors associated with TB screening and test ordering. Alt Text. Table providing the results of the multivariable logistic regression analysis for factors associated with TB risk factor screening and testing. Overall, younger children were more likely to be screened but less likely to be tested, and those who preferred a language other than English were more likely to be screened and tested.

| Variable | Assessment of TB Risk Factor<br>aOR (95% CI) | Testing for TB Infection<br>aOR (95% CI) |
| --- | --- | --- |
| **Age at visit in years** |  |  |
| Age 1 to 4 | REF | REF |
| Age 5 to 11 | 0.49 (0.41 – 0.58)** | 1.06 (0.72 – 1.55) |
| Age ≥12 years | 0.13 (0.11 – 0.15)** | 1.77 (1.20 – 2.63)<sup>1</sup> |
| **Sex** |  |  |
| Female | REF | REF |
| Male | 1.02 (0.92 – 1.14) | 0.74 (0.55 – 1.01) |
| **Race** |  |  |
| Asian | 1.03 (0.70 – 1.52) | 0.27 (0.06 – 1.19) |
| Black/African American | 0.97 (0.72 – 1.31) | 0.82 (0.32 – 2.08) |
| Muti-Racial | 1.25 (0.79 – 1.99) | 1.64 (0.58 – 4.65) |
| Native American /Alaskan Native | 1.36 (0.17 – 10.86) | - |
| Other | 0.99 (0.73 – 1.35) | 0.50 (0.20 – 1.23) |
| Pacific Islander/Native Hawaiian | 0.61 (0.33 – 1.13) | - |
| White/Caucasian | REF | REF |
| **Preferred Language** |  |  |
| English | REF | REF |
| Spanish | 1.25 (1.03 – 1.51)* | 1.67 (1.09 – 2.55)<sup>2</sup> |
| Arabic | 1.50 (1.03 – 2.17)* | 2.53 (1.46 – 4.40)<sup>1</sup> |
| Tigrinya/Tigre | 2.13 (1.13 – 4.03)* | 2.51 (1.38 – 4.56)<sup>1</sup> |
| Other | 1.15 (0.84 – 1.56) | 1.45 (0.88 – 2.40) |
| **TB Risk Factor** |  |  |
| Birth |  | REF |
| Travel/Residence |  | 0.76 (0.52 – 1.11) |
| Exposure |  | 0.69 (0.35 – 1.36) |
| “Risk” |  | 0.22 (0.12 – 0.41)<sup>1</sup> |
| Unknown |  | 0.35 (0.21 – 0.57)<sup>1</sup> |

aOR: Adjusted Odds Ratio; CI: Confidence Interval
1. $p < 0.01$
2. $p < 0.05$

## DISCUSSION

Despite the AAP recommendation for annual risk factor screening for TB infection in children in the US, there is limited data on the pediatric LTBI care cascade and interventions to support primary care providers. At a pediatric FQHC in a California county with high TB incidence, we found that risk factor screening was high, but only 20% of those with a risk factor were tested for TB infection. Younger children were more likely to be screened than older children, but less likely to be tested, and non-English speakers were more likely to be screened and tested. Notably, the incorporation of TB risk factor questions into the well-child and -adolescent EHR notes was associated with a significant and sustained increase in TB risk factor screening to >90%, and testing increased over time. Our findings suggest that ongoing gaps in the LTBI care cascade for children and adolescents could lead to underdiagnosis and treatment, but simple EHR-based interventions have the potential to improve TB care.

One of the strengths of this study is its focus on early steps in the pediatric LTBI care cascade, namely identifying patients that need testing through screening for risk factors. The majority of studies on LTBI care cascades started with diagnostic test ordered, as data on the population screened is not available.^17^ One review and meta-analysis of pediatric LTBI care cascade research identified only 7 out of 146 total studies where the entire care cascade was included, of which only two were conducted within a US patient population and neither study was based in a primary care setting.^11,18^ Our results show that capturing early steps is important, as we found gaps and differences based on age group and language preference that can guide future interventions.

Our finding of low testing is consistent with past work on the pediatric LTBI care cascade in the US. A retrospective cohort study at a primary care clinic in Nashville, Tennessee, which included 8,756 children with TB risk factors found that only 38% of children and adolescents < 18 years with a TB risk factor had a TST placed.^19^ A retrospective cohort study conducted at a community health center in Northern California serving primarily non-US born patients found that in 2019, 43% of patients with TB risk factors and no prior testing received a TB test.

After testing, most of those with a positive test were appropriately evaluated and, if indicated, initiated on treatment. A retrospective study in Boston, Massachusetts, similarly found that most pediatric patients with a positive test were evaluated (94%) and had a treatment recommendation (90%).^11^ However, we only identified nine children with positive TB diagnostic testing over the seven year period. Given the low proportion of patients with testing among those with TB risk factors, this may be an underestimate of the true TB infection prevalence and highlights the need to better understand and intervene on actionable barriers to testing. Only two-thirds of patients completed treatment, which may represent a limitation of documentation, but there are also known challenges in completion of LTBI treatment.^21^ The study in Boston similarly found only 59% of children with LTBI completed treatment or were recommended to not be treated.^11^

We found that gaps in the care cascade were associated with age group; adolescents 12 years and older had screening for risk factor rates below 90% during the entire study period, while younger patients under five years were less likely to have an LTBI test ordered. While the reasons for these differences require further investigation, young children have more frequent well-child visits than adolescents and thus there may be more opportunities to complete a risk factor screening. Although the study in Tennessee found that older children were less likely to be tested, the absolute age difference was small (median age 6 vs. 5.3 years) and they relied on TST.^19^ In our study, most were tested with an IGRA; this has the advantage of not requiring a follow up visit, but has the challenge of requiring venipuncture, which can be particularly challenging in younger children and may limit uptake in this age group. This was also found in the study in Boston, where adolescents were more likely to complete testing.^11^ Other factors could include reluctance in initiating treatment in younger children, perceptions of tuberculosis in family and providers, or lack of comfort in testing or treatment by providers.^21,22^

We found that patients who spoke a language other than English were more likely to be screened for TB risk factors, and have a test ordered if a risk factor was present. The study in Boston also found that non-English/non-Spanish speakers were more likely to complete testing.^11^ Non-English speaking may be used as a proxy for non-US birth and prompt providers to complete TB screening and testing. However, as a standalone approach, this could result in children being missed without standard risk factor assessment.

The addition of TB risk factor screening questions to the well-child and -adolescent notes was associated with a sustained increase in risk factor screening. We also saw an increase in TB infection testing, which may be due to greater provider awareness after completing TB risk factor screening. However, there may have been also other contributing factors including increased provider education and promotion of QFT testing in the clinic. The study in Tennessee also introduced a template in their EHR and similarly found that 96% of children completed screening. The prior study in Northern California also found an increase in testing rates from 23% to 80% by using a range of approaches that included TB-related diagnostic order sets, provider progress reports, and guided note templates.^20^ Significant data already exists for utilization of other EHR based interventions, such as nudges or best practice reminders, to increase rates of cancer screening and vaccination.^23–25^ Overall, this is one of the first studies to signal that an EHR-based intervention can increase TB infection screening for children in pediatric primary care, and suggests that additional tools can be introduced to improve other gaps in the care cascade, such as testing and documentation of treatment completion.

Our study benefited from utilizing a large number of well-child visits in an FQHC over a seven-year period in a county with a high TB incidence relative to the rest of the country. However, there are several limitations to our study. Building care cascades based on EHR data has challenges, including missing or incomplete information. Ongoing gaps in risk factor screening and testing may be related to past assessments that were not documented or not included in the extracted note. Moreover, TST or IGRA testing performed at another facility or as part of a public health investigation is not always entered into the EHR. We also did not have documentation if a test was refused by the patient or family. We focused on well-child and - adolescent visits to align with AAP screening recommendations and assessment among asymptomatic children, but TB risk assessment may have also occurred at “sick” visits or as part of requirements for schools and other organizations. The low numbers of patients with positive LTBI screening tests prevented analysis of factors associated with completion of later steps of the pediatric LTBI care cascade, although prior meta-analysis data suggests that the majority of patients who initiate LTBI treatment complete therapy.^26^ Lastly, the study was conducted at facilities that predominantly serve a population with public insurance. While the care cascade will vary in different facilities and settings, we demonstrated how the EHR can be used to develop care cascades and identify ongoing gaps.

## CONCLUSION

Overall, we found that gaps remain in the pediatric LTBI care cascade that could lead to underdiagnosis and missed treatment opportunities. The barriers to completing these steps are likely multi-factorial, and require additional exploration with providers, patients, and families to guide future interventions to improve the quality of care. At the same time, we demonstrated that simple, low-cost changes in the EHR such as a modification of a note template can lead to substantial improvements in risk factor screening, which may in turn improve completion of downstream steps. This supports that EHR-based tools should be considered as part of an intervention package to enable providers and families to address ongoing gaps in LTBI care for children and adolescents.

## Data Availability

All data produced in the present study are available upon reasonable request to the authors.

## Conflict of Interest Disclosures (includes financial disclosures)

The authors have no conflicts of interest to disclose.

## Role of Funder/Sponsor (if any)

The funder had no role in the design and conduct of the study.

## Clinical Trial Registration (if any)

Not applicable

## Abbreviations

AAP: American Academy of Pediatrics
aOR: adjusted odds ratio
BCG: Bacille Calmette-Guérin
CA: California
CDPH: California Department of Public Health
CI: confidence interval
EHR: electronic health record
FQHC: Federally Qualified Health Center
ICD: International Classification of Diseases
ICD-9: International Classification of Diseases, Ninth Revision
ICD-10: International Classification of Diseases, Tenth Revision
IGRA: interferon gamma release assay
IRB: Institutional Review Board
IQR: interquartile range
LTBI: latent tuberculosis infection
QFT: QuantiFERON-TB Gold Plus
TB: tuberculosis
TST: tuberculin skin testing
US: United States
WI: Wisconsin

## Contributors Statement

Julia Fink conceptualized the study, conducted the analysis and drafted the manuscript.

William Burrough conceptualized the study, collected the data, performed chart review, and edited the manuscript.

Mariamawit Tamerat guided the data collection, oversaw the implementation of the electronic health record TB screening, and edited the manuscript.

Shereen Katrak, Tessa Mochizuki, Katya Salcedo, Amit Chitnis, and Charlotte Hsieh, oversaw the data collection and analysis, guided interpretation, and edited the manuscript.

Zarin Noor and Gena Lewis conceptualized the study, oversaw the implementation of the electronic health record TB screening, guided data collection and interpretation, and edited the manuscript.

Devan Jaganath conceptualized the study, obtained resources, guided analyses and interpretation, and drafted and edited the manuscript.

All authors approved the final manuscript as submitted and agree to be accountable for all aspects of the work.

