## Supplemental for "Latent tuberculosis infection diagnosis and management in pediatric primary care: The role of electronic health record-based screening"

**SUPPLEMENTAL MATERIAL**

**Supplemental Methods. Definition of LTBI Care Cascade Steps**

*LTBI Risk Factor Screening*

Completion of risk factor screening was defined if a provider indicated “yes”, “no” or selected a risk factor. Incompletion was defined if no response was included after a risk factor screening question or a selection was not made for the drop-down smart list.

*Positive LTBI Risk Factor*

For patient visits with risk factor screening, a positive risk factor screening assessment was defined by “yes” or a smart list selection of a LTBI risk factor. Any explanatory free text was reviewed to guide the reason for a positive screen.

*Risk Factor Classification*

Risk factors were categorized as: 1) Birth outside US; 2) Travel of greater than one month to a region with endemic TB; 3) Exposure to TB; 4) Other risk factor; and 5) Not classified. The exposure to TB risk group included children where the provider documented exposure to TB disease or LTBI. While LTBI contact is not included in the CDPH Pediatric TB Risk Assessment, we included this factor as the AAP recommends testing for recent household conversion of TB infection and to reflect provider ordering.[1] The other risk factor group represented patients who providers determined had risk factors for TB exposure due to living conditions or situations other than risk factors defined by the CDPH Pediatric TB Risk Assessment. These patients included those living in group homes, recently incarcerated, unhoused, living in shelters, or living with someone who was recently incarcerated/unhoused. Not classified indicates no risk factor for TB disease was reported.

*LTBI Diagnostic Test Ordering*

LTBI diagnostic test options included a tuberculin skin test (TST) or QuantiFERON TB Gold-In-Tube or Plus; (QFT; Qiagen; Germantown, Maryland, USA). To determine if an LTBI diagnostic test was ordered, we analyzed visit-specific diagnosis and visit-specific orders for patients with a provider-identified TB risk factor. Patients were considered to have TB test ordered if the encounter order included “read PPD”, “TB Skin Test” or “QuantiFERON Gold test.” Patients were also considered to have a screening test ordered if their encounter diagnosis included TB, Tuberculosis or PPD as per clinic protocol. We included all tests that were ordered during or after the analyzed visit but prior to the next documented patient clinic visit. Patients were categorized as having a PPD ordered if their encounter order was “read PPD” or “TB Skin Test” or if their diagnostic test information was obtained through encounter diagnosis. Patients were categorized as having a QFT ordered if their encounter order was “QuantiFERON Gold test.”

*Completion of LTBI Diagnostic Test*

Test results were extracted from the EHR, and chart review for nursing notes documenting TB test results was performed for those with missing test results. If either of these were found, a patient was considered to have an LTBI diagnostic test completed.

*Chest Imaging Ordering and Completion*

Chest x-ray (CXR) order status was extracted from patient data via chart review. CXR completion and results were extracted via chart review.

*LTBI Diagnosis*

Patients were given an LTBI diagnosis for analysis if they had a positive LTBI diagnostic test (TST or QFT), were asymptomatic, and had a CXR without evidence of TB disease as determined through provider note review and prescription initiation.[2]

*LTBI Treatment Initiation*

LTBI regimens for this cohort included a 9-month course of Isoniazid (INH) or a 4 month course of Rifampin (RIF). A patient was considered to have TB treatment initiated if either of these medications were ordered with progress notes that indicated LTBI treatment was started and/or continued.

*LTBI Treatment Completion*

Completion of LTBI treatment was determined by documentation of treatment completion in a follow-up visit note.

**Supplemental Table 1. Encounter Diagnosis Codes and ICD 9 and ICD 10 codes used for inclusion in analysis.**

| | |
|---|---|
| **Encounter Diagnoses** | *Well-Child/adolescent/adult, Routine child health, Health check, general adult medical, School physical/health, Sports physical/routine sports, Kindergarten physical for school admission, Encounter for school history and physical exam, Encounter for health supervision*, and *Care of other healthy infant and child* |
| **ICD 9 and 10 Codes** | A12.81-85, A15.X, A17.0, A17.1, A17.8, A17.81-89, A17.9, A18.01-03, A18.09, A18.10-18, A18.31, A18.32, A18.39, A18.4, A18.50-54, A18.59, A18.6, A18.7, A18.89, A19.0-2, A19.8, A19.9, Z22.7, R76.11-2, Z20.1, Z86.11 Z91.89, Z92.89, 011.0–018.96 |

**Supplemental Table 2. Univariate factors associated with TB risk factor screening at visit-level**

| Variable | TB Risk Factor Assessment (n = 18,681)[1] | | p-value |
|---|---|---|---|
| | **Risk Assessed (n= 16,838)** | **Not Assessed (n=1,843)** | |
| **Age at visit in years** (median, IQR) | 6.3 (2.9, 11.3) | 12.5 (7.9, 14.7) | |
| **Age at visit in years** (N, %) | | | <0.01 |
| Age 1 to 4 | 6,820 (96.5) | 251 (3.6) | |
| Age 5 to 11 | 6,445 (92.6) | 515 (7.4) | |
| Age 12+ | 3,573 (76.8) | 1,077 (23.2) | |
| **Sex** (N, %) | | | 0.36 |
| Female | 8,373 (49.7) | 937 (50.8) | |
| **Race** (N, %) | (n=15,177) | (n=1,650) | 0.02 |
| Asian | 784 (5.2) | 73 (4.4) | |
| Black/African American | 6,492 (42.8) | 760 (46.1) | |
| Muti-Racial | 466 (3.1) | 33 (2.0) | |
| Native American /Alaskan Native | 20 (0.1) | 1 (0.1) | |
| Other | 6,638 (43.7) | 691 (41.9) | |
| Pacific Islander/Native Hawaiian | 99 (0.7) | 15 (0.9) | |
| White/Caucasian | 678 (4.5) | 77 (4.7) | |
| **Ethnicity**[2] (N, %) | (n=15,335) | (n=1,667) | 0.16 |
| Hispanic | 3,800 (24.8) | 387 (23.2) | |
| **Preferred Language** (N, %) | (n=16,836) | (n=1,843) | <0.01 |
| English | 13,569 (80.6) | 1,554 (84.3) | |
| Spanish | 1,866 (11.1) | 190 (10.3) | |
| Arabic | 515 (3.1) | 34 (1.8) | |
| Tigrinya/Tigre | 262 (1.6) | 14 (0.8) | |
| Other | 624 (3.7) | 51 (2.8) | |
| **Preferred Language** (N, %) | (n=16,836) | (n=1,843) | <0.01 |
| Non-English | 3,267 (19.4) | 289 (15.7) | |
| **Non-English Speakers** (N, %) | (n=3,267) | (n=289) | <0.01 |
| Spanish | 1,866 (57.1) | 190 (65.7) | |
| Other | 1,401 (42.9) | 99 (34.3) | |
| **Insurance Type** (N, %) | (n=16,786) | (n=1,837) | 0.35 |
| Public | 15,505 (92.4) | 1,708 (93.0) | |

IQR – interquartile range

1. The denominator is the column total unless otherwise noted.
2. Not-imputed, only analyzed patient if selected “Hispanic or Latino” or “Not Hispanic or Latino”

**Supplemental Table 3: Univariate factors associated with TB infection testing at visit-level (n=1,277 visits)**

| Variable | Total Unique Patients (n=1,165) | TB Infection Test Ordered[1] (n= 261 visits) | No TB Infection Test Ordered[1] (n=1,016 visits) | p-value |
|---|---|---|---|---|
| **Age at visit in years** (median, IQR) | 7.1 (3.6, 12.8) | 9.1 (4.4, 14.7) | 6.7 (3.4, 12.0) | <0.01 |
| **Age at visit in years** (N, %) | | | | <0.01 |
| Age 1 to 4 | 415 (35.6) | 75 (28.7) | 388 (38.2) | |
| Age 5 to 11 | 419 (36.0) | 86 (33.0) | 373 (36.7) | |
| Age 12+ | 331 (28.4) | 100 (38.3) | 255 (25.1) | |
| **Sex** (N, %) | | | | 0.58 |
| Female | 593 (50.9) | 137 (52.5) | 514 (50.6) | |
| **Race** (N, %) | (n = 1,009) | (n=213) | (n=897) | <0.01 |
| Asian | 93 (9.2) | 21 (8.1) | 87 (8.6) | |
| Black/African American | 276 (27.3) | 45 (17.2) | 254 (25.0) | |
| Muti-Racial | 21 (2.1) | 4 (1.5) | 18 (1.8) | |
| Native American /Alaskan | 2 (0.2) | 0 (0) | 3 (0.3) | |
| Native | 582 (57.7) | 132 (50.6) | 510 (50.2) | |
| Other | 3 (0.3) | 3 (1.2) | 0 (0) | |
| Pacific Islander/Native | 32 (3.2) | 8 (3.1) | 25 (2.5) | |
| Hawaiian | | | | |
| White/Caucasian | | | | |
| **Ethnicity[2]** (N, %) | (n = 1,028) | (n=213) | (n=917) | 0.32 |
| Hispanic | 302 (29.4) | 68 (31.9) | 261 (28.5) | |
| **Preferred Language** (N, %) | (n = 1,164) | (n=260) | (n=1016) | <0.01 |
| English | 719 (61.8) | 114 (43.9) | 672 (66.1) | |
| Spanish | 190 (16.3) | 60 (23.1) | 146 (14.4) | |
| Arabic | 79 (6.8) | 30 (11.5) | 56 (5.5) | |
| Tigrinya/Tigre | 59 (5.1) | 23 (8.9) | 41 (4.0) | |
| Other | 117 (10.1) | 33 (12.7) | 101 (9.9) | |
| **Preferred Language** (N, %) | (n = 1,164) | (n=260) | (n=1016) | <0.01 |
| Non-English | 445 (38.2) | 146 (56.2) | 344 (33.9) | |
| **Non-English Speakers** (N, | (n = 445) | (n=146) | (n=344) | 0.78 |
| %) | 190 (42.7) | 60 (41.1) | 146 (42.4) | |
| Spanish | 255 (57.3) | 86 (58.9) | 198 (57.6) | |
| Other | | | | |
| **Insurance Type** (N, %) | (n = 1,155) | (n=328) | (n=939) | 0.23 |
| Public | 1089 (94.2) | 246 (95.7) | 947 (93.8) | |
| **TB Risk Factor** (N, %) | | | | <0.01 |
| Birth | 355 (30.5) | 130 (49.8) | 261 (25.7) | |
| Travel/Residence | 298 (25.6) | 73 (28.0) | 268 (25.2) | |
| Exposure | 76 (6.5) | 14 (5.4) | 63 (6.2) | |
| Immunocompromised | 4 (0.3) | 0 (0) | 5 (0.5) | |
| “Risk” | 222 (19.0) | 18 (6.9) | 222 (21.9) | |
| Unknown | 210 (18.0) | 26 (10.0) | 197 (19.4) | |

IQR – interquartile range

1. The denominator is the column total unless otherwise noted.
2. Not-imputed, only analyzed patient if selected “Hispanic or Latino” or “Not Hispanic or Latino”

**Supplemental Figure 1. Care cascade for pediatric TB infection among children under 5 years old.** Each bar represents the total number of visits where the step was completed or result was positive, and is the denominator for the next step. The pie chart above each bar indicates the proportion completed or positive.

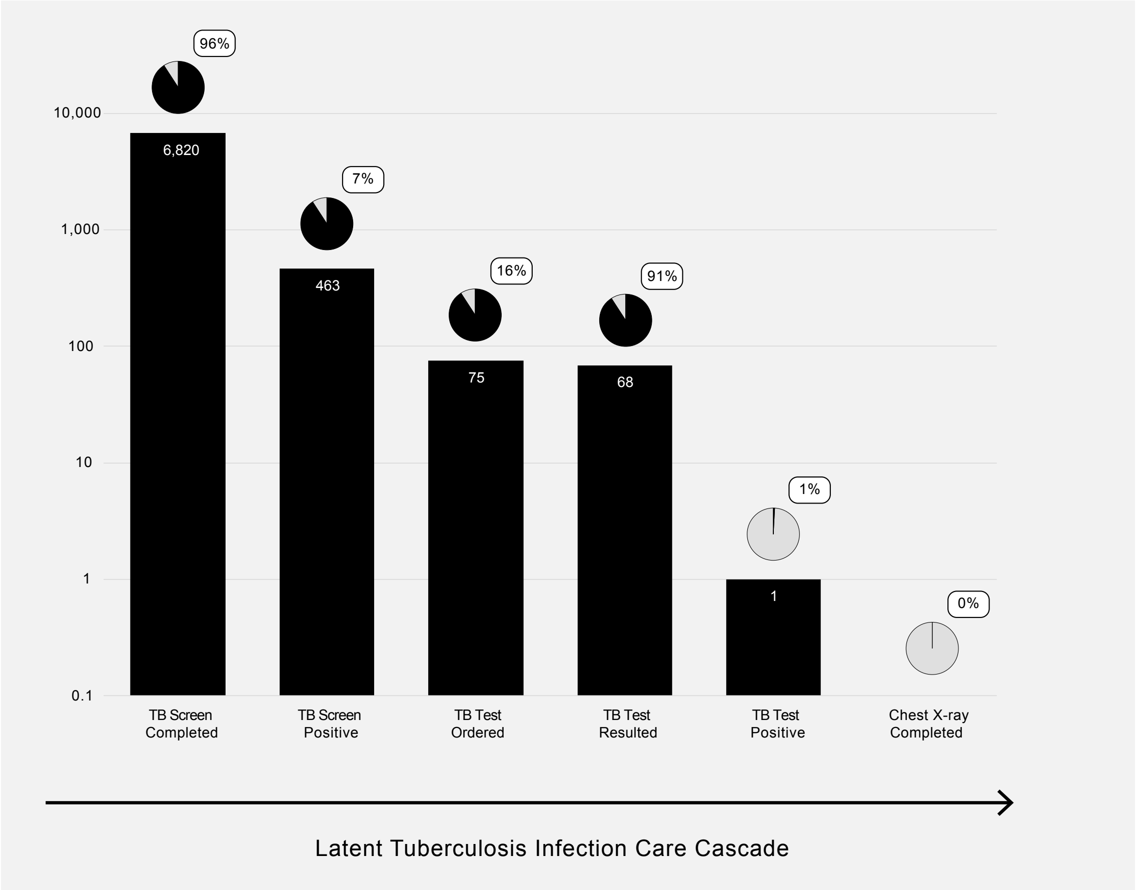

**Supplemental Figure 2. Care cascade for pediatric TB infection among children 5-11 years.** Each bar represents the total number of visits where the step was completed or result was positive, and is the denominator for the next step. The pie chart above each bar indicates the proportion completed or positive.

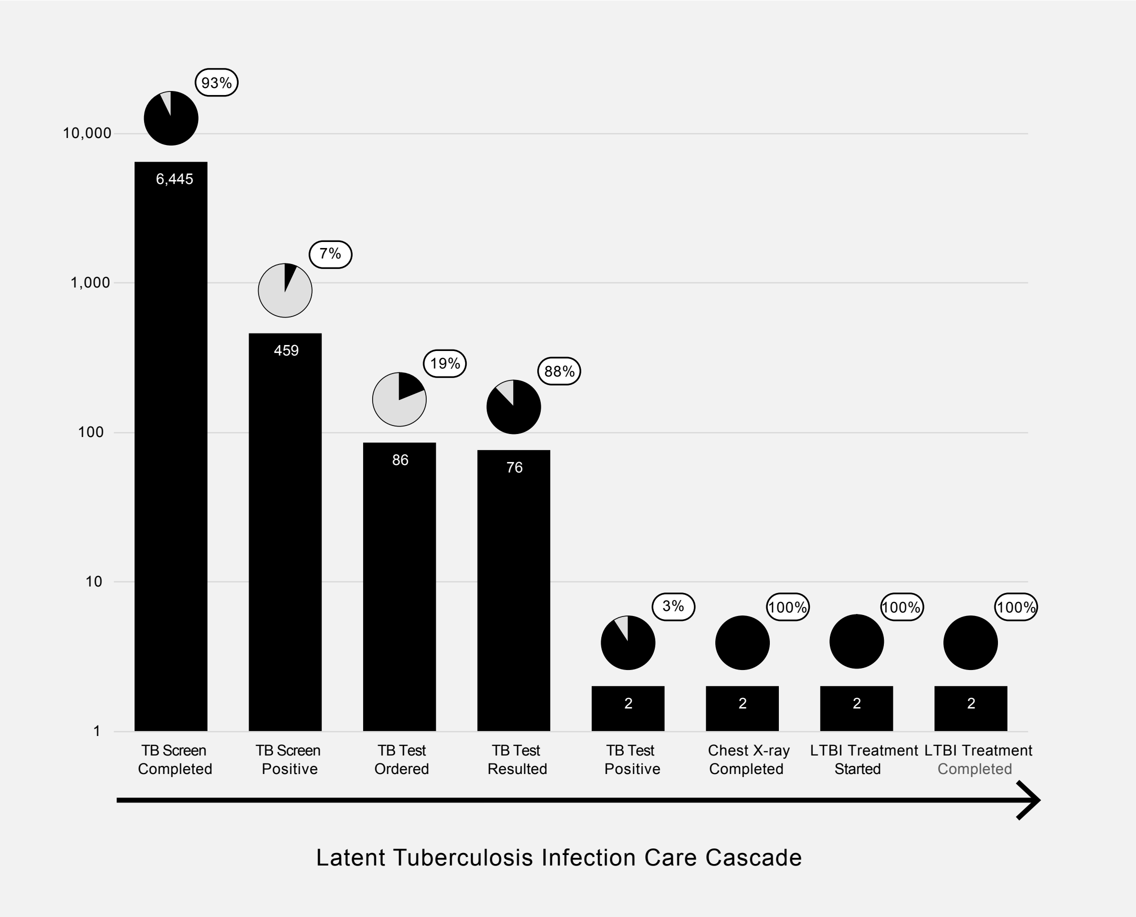

**Supplemental Figure 3. Care cascade for pediatric TB infection among children 12 years and older.** Each bar represents the total number of visits where the step was completed or result was positive, and is the denominator for the next step. The pie chart above each bar indicates the proportion completed or positive.

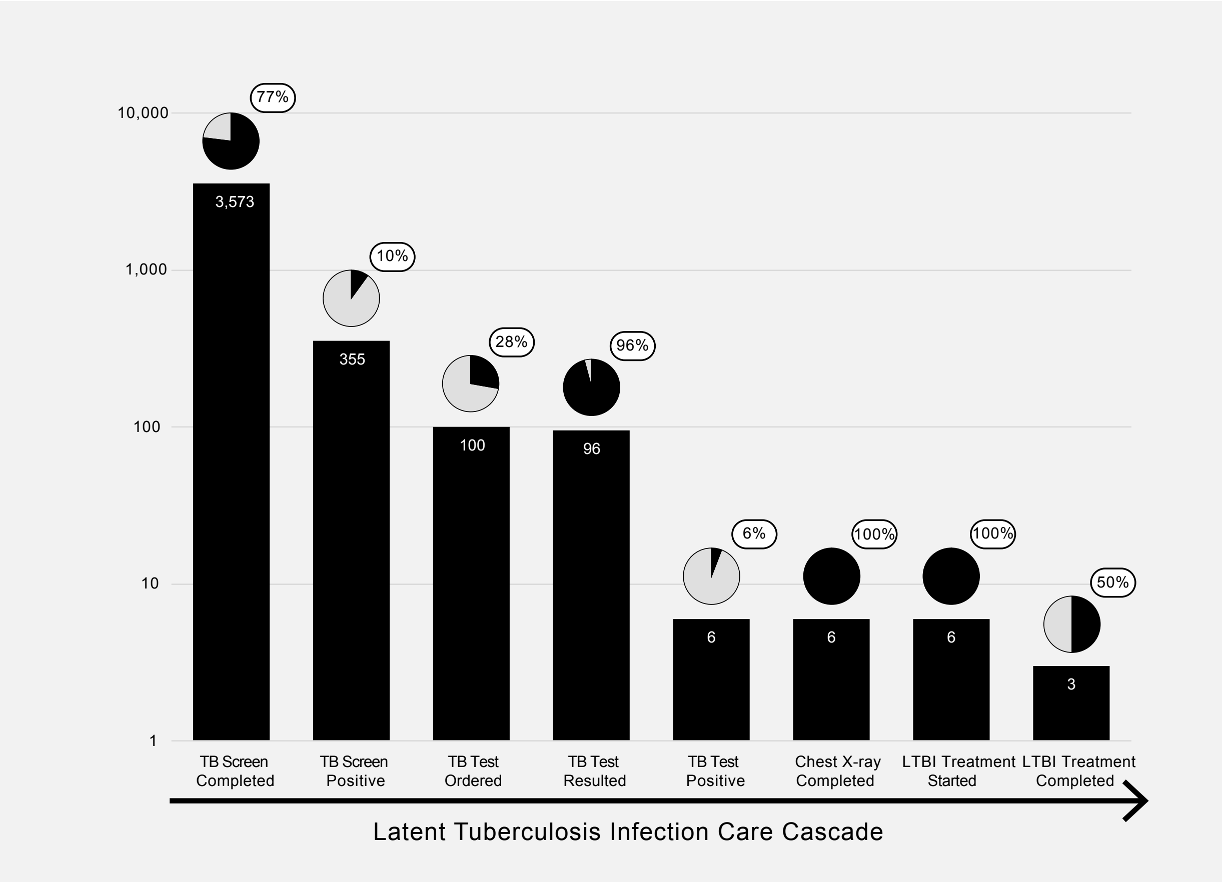

**Supplemental Figure 4. Care cascade for pediatric TB infection among children who prefer a language other than English.** Each bar represents the total number of visits where the step was completed or result was positive, and is the denominator for the next step. The pie chart above each bar indicates the proportion completed or positive.

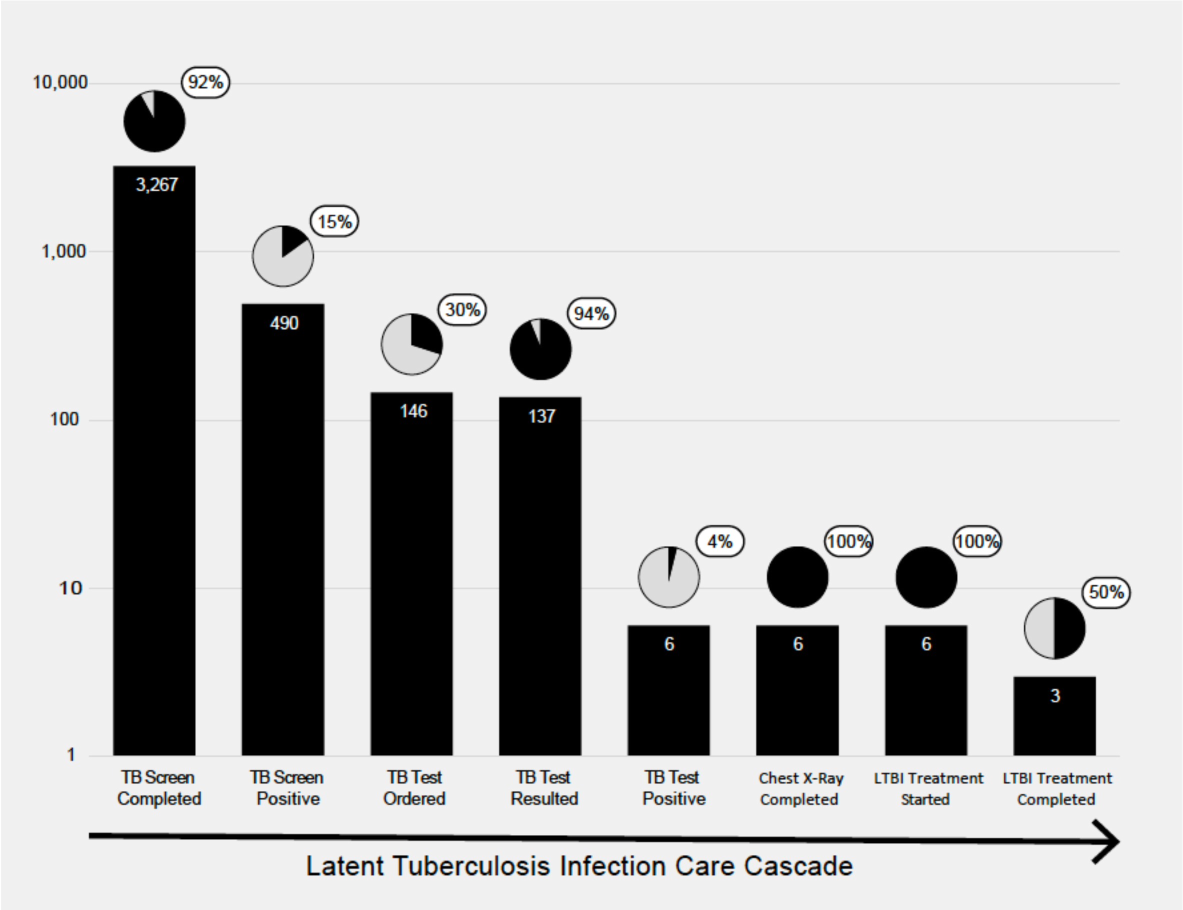

**Supplemental Figure 5. Annual TB risk factor screening by age category.** Percentage TB Screening in well visits by patient age categories, and overall screening rate for cohort.

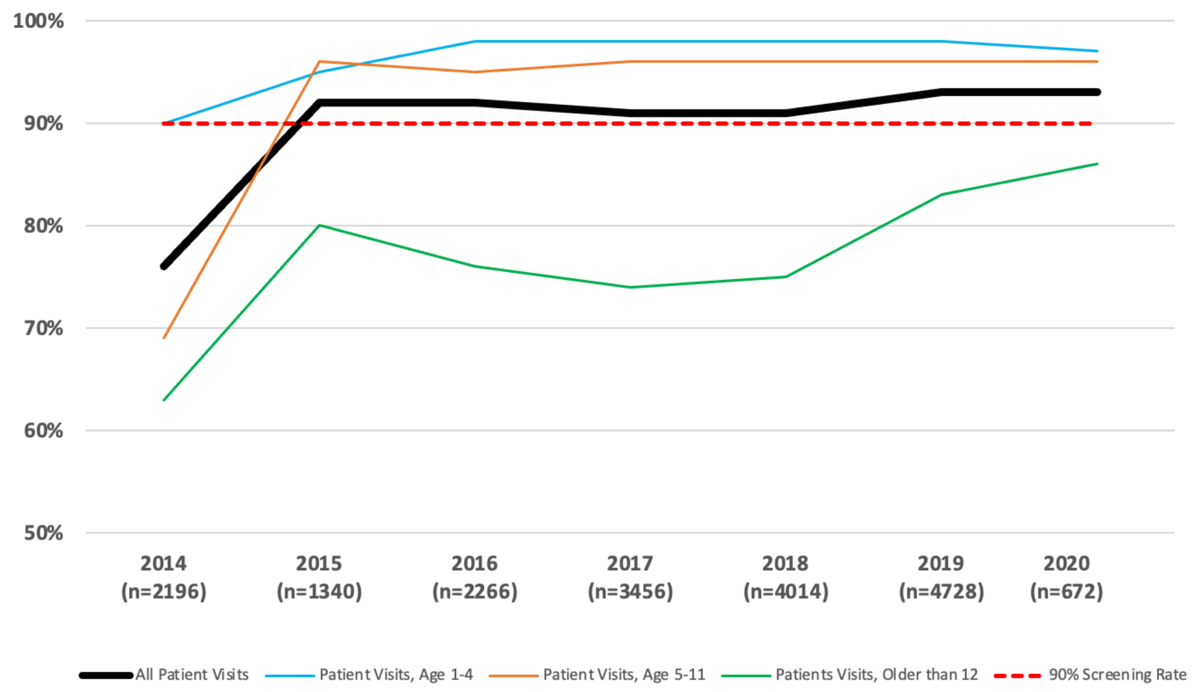

**Supplemental Figure 6. Annual proportion of TB infection testing by age group.** TB testing included tuberculin skin test or QuantiFERON Gold, and only included children with a TB risk factor.

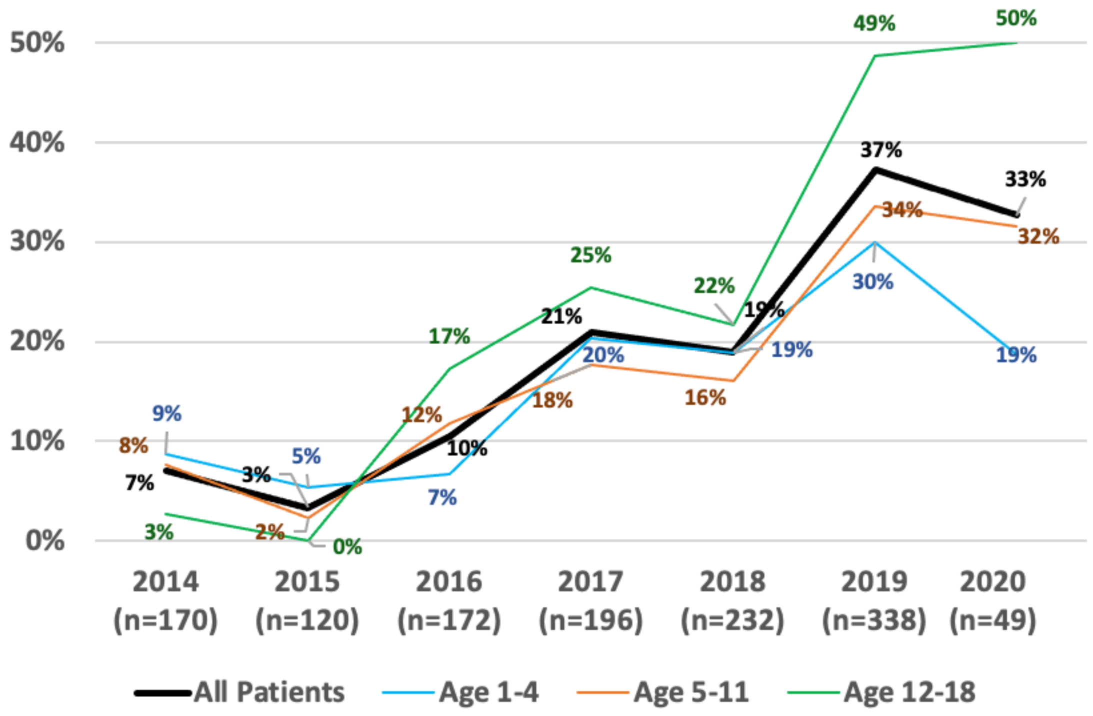

135

---

[1] American Academy of Pediatrics. RED BOOK : report of the committee on infectious diseases. Elk Grove Village: Amer Acad Of Pediatrics; 2024.

[2] CDPH CTCA JOINT GUIDELINE 2018 REVISION [Internet]. [cited 2025 Jan 28]. Available from: https://ctca.org/wp-content/uploads/2018/12/LTBI-Guidelines-2018-Revision-FINAL.pdf\
